# Hippocampal microstructural and neurobehavioral differences in welders are related to higher R2* in the red nucleus

**DOI:** 10.1101/2022.06.03.22275885

**Authors:** Eun-Young Lee, Juhee Kim, Janina Manzieri Prado-Rico, Guangwei Du, Mechelle M. Lewis, Lan Kong, Jeff D. Yanosky, Byoung-Gwon Kim, Young-Seoub Hong, Richard B. Mailman, Xuemei Huang

## Abstract

**Introduction:** Metal exposure has been associated with higher risk of neurodegenerative disorders such as Alzheimer’s disease (AD). We examined the potential link between welding-related metal co-exposure (e.g., Fe, Mn, Pb) and AD-related structural and neurobehavioral metrics.

**Methods:** Subjects with (welders; n=42) or without (controls; n=31) a history of welding were examined. Metal exposure was estimated by exposure questionnaires and whole blood metal levels. Brain metal accumulations were estimated by MRI R1 (Mn) and R2* (Fe) in the caudate, putamen, globus pallidus, red nucleus (RN), and hippocampus. AD-related structural differences were assessed by volume and diffusion tensor imaging metrics in the hippocampus, and neurobehavioral aspects by learning/memory task scores.

**Results:** Compared to controls, welders displayed higher blood metal levels (p’s <0.004) and R2* values in the caudate and RN (p’s<0.024). Caudate R2* values were associated with blood Fe (p=0.043), whereas RN R2* values were correlated with blood Pb (p=0.003). Welders had higher hippocampal mean diffusivity (MD; p=0.011) and lower *Story Recall* scores (p=0.049), but no difference in volume or domain-wise learning/memory performance (p’s>0.117). Group differences in hippocampal MD and *Story Recall* scores were greater with higher RN R2* values (p’s<0.016). Moreover, RN R2* values reflected an indirect link between blood Pb and hippocampal MD (p=0.036) across both groups.

**Discussion:** Welders had hippocampal structural and learning/memory performance differences similar to those in AD-at-risk populations. These AD-like differences in welders may, in part, be linked to Pb exposure reflected by higher RN R2* levels at the brain level.

## Introduction

There are growing concerns about health consequences of exposure to toxicants found in vehicle emissions, drinking water, and occupation-related activities. Many of these compounds contain metals (e.g., Fe, Mn, and Pb) that can cross the blood-brain barrier, accumulate in specific brain regions, affect cellular functions, and exert neurotoxic effects (Teleanu et al. 2018; Zhou et al. 2018). Previous studies suggested that chronic exposure to metals is associated with brain changes that may lead to neurodegenerative disorders (Bakulski et al. 2020; Huat et al. 2019). Even essential elements (e.g., Fe and Mn) can be neurotoxic at high doses (Dobson et al. 2004; Salvador et al. 2011). In contrast, Pb is a non-essential element and known to cause neurotoxic effects even at very low exposure levels (Alves Oliveira et al. 2020).

Alzheimer’s disease (AD) is the most common age-related neurodegenerative disorder, comprising about 70% of dementia cases (Brookmeyer et al. 2011). It is characterized by accumulations of β-amyloid plaques and tau tangles in the brain, with the most prominent neuronal damage noted in the hippocampus (Li et al. 2019; Sørensen et al. 2016). The major behavioral deficits entail learning/memory problems (Bäckman et al. 2005). Once AD-related clinical symptoms are manifested, the disease progresses relentlessly (Li et al. 2019). Given the widespread nature of exposure to metals in the general population, potential links between exposure to metals and AD have been sought. Indeed, studies in metal-exposed animals reported higher accumulation of these metals that was associated with AD-like Aβ production and cognitive impairment (Guilarte 2010; Schneider et al. 2013; Schroeder et al. 2013). Epidemiological studies have found higher plasma levels of these metals in AD populations (Gerhardsson et al. 2008; Hare et al. 2016; Vural et al. 2010).

Linking chronic life-long metal exposure to age-related neurodegenerative diseases in humans, however, is challenging due to the long-latency of such diseases and the possibility that early disease-related changes may be subtle, proceed unnoticed, or be blamed on normal aging processes. Thus, there is increased interest in detecting biomarkers that capture AD-related early brain changes. For example, MRI T1-weighted images have been utilized to measure cortical thinning and hippocampal volume to estimate AD-related neuronal death (Koval et al. 2021; McRae-McKee et al. 2019). Recent studies suggested that diffusion tensor imaging (DTI) metrics such as higher mean diffusivity (MD) and lower fractional anisotropy (FA), measuring random translational water motion affected by microstructural changes, may be more sensitive than hippocampal volume in capturing AD-related early brain changes (Douaud et al. 2013; Fellgiebel et al. 2006; Müller et al. 2007). As a result, these DTI metrics have been hypothesized to estimate disease-related early microstructural brain tissue changes (Le Bihan et al. 2001).

Welding fumes are enriched in various metals (e.g., Fe, Mn, Pb) (Abdullahi and Sani 2020). Previous welding-related studies, however, have focused mainly on the neurotoxic effects of excessive Mn exposure in the basal ganglia (BG) and its associations with parkinsonism-related neurobehavioral symptoms and disease development (Colosimo and Guidi 2009; Guilarte and Gonzales 2015). More recent studies have demonstrated that Mn accumulation occurs in brain areas separate from the BG, including the hippocampus (Lee et al. 2015b; Liang et al. 2015). Moreover, metals other than Mn (e.g., Fe or Pb) may accumulate in specific brain regions [e.g., frontal cortex (Long et al. 2014), caudate nucleus (CN) (Lee et al. 2016), and red nucleus (RN) (Prado-Rico et al. 2022)] when welding exposure is chronic but relatively low. Recently, we reported higher hippocampal MD values in welders (Lee et al. 2019), structural differences similar to those seen in AD-at-risk populations. Of particular interest is the potential association of welding-related metal co-exposure and these AD-related metrics, and possible neurodegenerative processes.

The present study examined the link between metal co-exposure and AD-related structural and neurobehavioral metrics in welders. Metal exposure was estimated by exposure questionnaires, whole blood metal levels, and brain metal accumulations measurements (MRI R1 for Mn and R2* for Fe) to reproduce higher metal exposure and brain metal accumulations in welders as reported previously (Prado-Rico et al. 2022). AD-related structural metrics were assessed by volume and DTI measures in the hippocampus, and AD-related neurobehavioral performance by learning/memory task scores. Our central hypotheses were: 1) hippocampal volume and FA will be lower and MD higher in welders; 2) welders will show lower performance on learning/memory tasks; 3) hippocampal structural metrics will be associated with learning/memory performance scores; 4) welding-related metal exposure will be associated with AD-related hippocampal structural and learning/memory task metrics.

## Methods

### Study subjects

Eighty subjects were recruited from labor unions in central Pennsylvania (USA) and the surrounding community. Welders were defined as subjects who had welded at any point in their lifetime, and controls as those without a history of welding. All subjects answered negatively for past Parkinson’s disease (PD) diagnosis or other neurological disorders, and were free of any obvious neurological or movement deficits using the Movement Disorders Unified PD Rating Scale motor exam subscore (UPDRS-III) with a threshold score of <15. Global cognition was measured by administering the Montreal Cognitive Assessment (MoCA). All subjects were male and non-demented [Mini-Mental Status Examination (MMSE) scores >24 or MoCA >19]. Demographic information, including age and education years, also was acquired. Forty-two welders and 31 controls completed the MRI acquisition with good quality images. Data from 2 controls and 7 welders were excluded for the R2* data analysis due to poor co-registration issues. This study was conducted at PennStateHealth (PSH) in compliance with the ethical principles of the Declaration of Helsinki and guidelines for Good Clinical Practice issued by the International Conference on Harmonization. It was reviewed and approved by the PSH Institutional Review Board.

### Overall exposure measurements

Metal exposure from welding fumes was measured on the same day using exposure questionnaires; whole blood metal levels of Fe, Mn, and Pb; and MRI R1 and R2* values (estimates of brain Mn and Fe accumulation, respectively; *vide infra* for details).

#### Exposure questionnaires

We utilized established exposure questions (Lee et al. 2015a) to estimate the following metrics: 1) recent exposure in the 90 days prior to the study visit including hours welding [HrsW90 = (weeks worked) * (h/week) * (fraction of time worked related directly to welding) and E90 (an estimate of the cumulative exposure to welding fumes)], and 2) lifetime exposure including welding years [(YrsW=years spent welding during the subjects’ life) and ELT (an estimate of cumulative exposure to welding fumes over the individual’s life)] (Lee et al. 2015a).

#### Blood analysis

To estimate whole blood metal levels, whole blood samples were stored at -80°C until analysis. Whole blood metal levels were analyzed by Inductively Coupled Plasma Mass Spectrometry (ICP-MS) (Lee et al. 2015a).

### MRI image acquisition and image processing

All MR images were acquired using a Siemens 3 T scanner (Magnetom Trio, Erlangen, Germany) with an 8-channel head coil. First, high-resolution T1-weighted (T1W) and T2-weighted (T2W) images were acquired for anatomical segmentation. For T1W images, MPRAGE sequences with Repetition Time (TR)/Echo Time (TE)=1540/2.3 ms, FoV/matrix=256×256/256×256 mm, slice thickness=1 mm, slice number=176 (with no gap), and voxel spacing 1×1×1 mm were used. T2W images were obtained using fast-spin-echo sequences with TR/TE=2500/316 ms and the same spatial resolution as the T1W images. For R1, TR/TE=15/1.45 ms, flip angles=4/25°, FoV/matrix=250×250/160×160 mm, slice thickness=1 mm, slice number=192, and voxel spacing=1.56×1.56×1 mm were used. R2* images were acquired using five TEs ranging from 8-40 ms with an interval of 8 ms, TR=51 ms, flip angle=15°, FoV/matrix=230×230/256×256 mm, slice thickness=1.6 mm, and slice number=88 were used. For DTI, TR/TE=8300/82 ms, b value=1000 s/mm^2^, diffusion gradient directions=42 and 7 b=0 scans, FoV/matrix=256×256/128×128 mm, slice thickness=2 mm, and slice number=65 were used.

### Defining brain regions of interest

Brain regions that previously had reported higher welding-related metal accumulation [bilateral caudate (CN), putamen (PUT), globus pallidus (GP), red nucleus (RN), and hippocampus (Figure 1) (Criswell et al. 2012; Lee et al. 2015a; Prado-Rico et al. 2022)] were selected as regions-of-interest (ROI). The ROIs were defined for each subject using automatic segmentation software (AutoSeg) or Freesurfer (for the hippocampus). The segmentation quality then was confirmed visually by a reviewer blinded to group assignment. The right and left hemisphere MRI data values were averaged.

**Figure 1.**
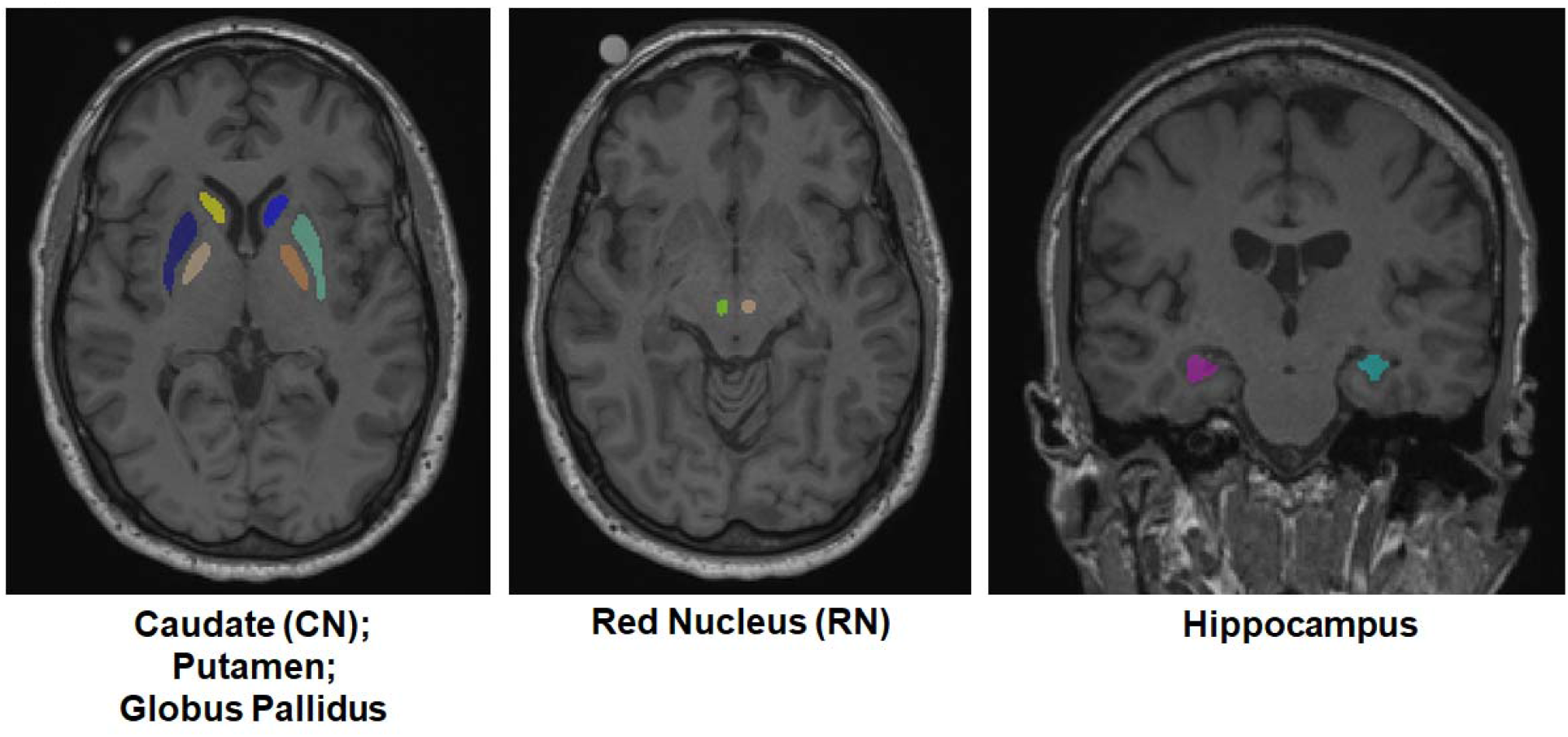
Automatically segmented regions of interest [caudate (CN), putamen, globus pallidus (GP), red nucleus (RN), and hippocampus] on T1-weighted MPRAGE images.

### Estimates of brain metal accumulation

#### MRI R1 values

To calculate R1 values (1/T1), ROIs were co-registered onto the whole brain T1 time images generated by the scanner using an affine registration implemented in 3D Slicer (www.slicer.org; Rueckert et al. 1999).

#### MRI R2*values

To calculate the apparent transverse relaxation rate (R2*=1/T2*), the magnitude images of multigradient-echo images were used. A voxel-wise linear least-squares fit to a mono-exponential function with free baseline using in-house Matlab (The MathWorks, Inc., Natick, MA) tools was utilized to generate R2* values. ROIs were co-registered onto the R2* maps using an affine registration implemented in 3D Slicer (www.slicer.org; Rueckert et al. 1999). MRI R1 and R2* values were calculated in each voxel and averaged over each ROI.

### AD-related hippocampal structural metrics

#### Volume

Hippocampal volumetric segmentation was performed with the Freesurfer image analysis suite (http://surfer.nmr.mgh.harvard.edu/). The processing included motion correction, removal of non-brain tissue using a hybrid watershed/surface deformation procedure (Ségonne et al. 2004), automated Talairach transformation, and segmentation of the deep gray matter volumetric structures including the hippocampus (Fischl et al. 2002; Fischl et al. 2004).

#### DTI metrics

DTI quality control and tensor reconstruction were performed using DTIPrep (University of North Carolina, Chapel Hill, NC) that first checks diffusion images for appropriate quality by calculating the inter-slice and inter-image intra-class correlation, and then corrects for the distortions induced by eddy currents and head motion. DTI maps then were estimated via weighted least squares. The DTI maps were co-registered onto T1/T2W images using ANTS, and the transformation matrix was applied inversely to bring the hippocampal region to the DTI maps. Two DTI indices [fractional anisotropy (FA) and mean diffusivity (MD)] were calculated out of three diffusivity eigenvalues (Le Bihan et al. 2001). FA is a weighted average of pairwise differences of the three eigenvalues and may represent the degree of diffusion anisotropy. MD is an average of the three eigenvalues, providing the overall diffusion magnitude (Le Bihan et al. 2001).

### AD-related neurobehavioral metric: learning/memory tasks

To assess AD-related neurobehavioral differences, performance on learning/memory tasks was measured by administering subtests from the *Repeatable Battery for the Assessment of Neuropsychological Status (RBANS) (Randolph et al. 1998)*. Specifically, the *Verbal List-Learning and Recall, Immediate Story Recall, Delayed Story Recall*, and *Figure Recall* subtests from the *RBANS* were used to assess learning/memory performance of verbal and visuospatial information that often is associated with AD-related neurobehavioral functional decline. For the *Verbal List Learning* subtest, a non-organized list of 10 words was presented orally and subjects immediately recalled the word list. Four learning trials were performed where subjects were encouraged to recall the words in any order, even if they had mentioned them in a previous learning trial. For the *Verbal List Recall* subtest, subjects recalled the same word list after a delay of approximately 20 minutes that reflects long-term verbal memory function. For the *Immediate Story* subtest, subjects were presented with a short prose passage and immediately reproduced the story they heard. For the *Story Recall* subtest, subjects recalled the story after a delay of approximately 20 minutes. For the *Figure Recall* subtest, subjects reproduced a figure after a ∼20 minute delay that reflects visual long-term memory. Individual raw subtest scores were converted to age-adjusted norm scores (e.g., T-scores) and subsequently to z-scores. An average z-score was derived from the individual subtests to create a summary learning/memory domain score.

### Statistical analysis

Group comparisons were conducted using one-way analysis of variance (ANOVA). For group comparisons of standard neuropsychological tests, age-adjusted norm scores were analyzed using analysis of covariance (ANCOVA) with adjustment for education level. For MRI measures, analysis of covariance (ANCOVA) was conducted with adjustment of age. For the MRI R1 and R2* measures, the analyses additionally were adjusted by R2* or R1 values, and whole blood metal levels of Fe (for the R1 analysis) or Mn (for the R2* analysis) due to a potential interaction between Mn and Fe. When comparing hippocampal volume, total intracranial volume (TIV) and education level additionally were used as covariates. For hippocampal DTI FA and MD values, education level was included as a covariate.

Association analyses were conducted for controls and welders separately using Pearson or Spearman (indicated when conducted) partial correlation analyses with adjustment for potential confounders. For the association analyses between MRI metrics (R1 and R2*) and exposure metrics (HrsW_90_, E_90_, YrsW, ELT), age-adjusted R1 and R2* values were used. For the association analyses between MRI metrics (R1 and R2*) and blood metal levels of interest, the analyses were adjusted for age and the other blood levels. For the association analyses between AD-related metrics (hippocampal and neurobehavioral learning/memory performance metrics) and exposure metrics, age-adjusted hippocampal metrics and neurobehavioral scores were used. The association analyses between AD-related metrics and blood metal levels of interest were adjusted for the other blood levels in addition to age. The association analyses among AD-related metrics were conducted with adjustment for age and education level.

In order to determine metal exposure measurement that could explain the variances of those AD-related metrics that showed significant group differences, a stepwise regression analysis was conducted for controls and welders separately, as well as collapsed across groups. For the stepwise regression analysis, exposure measurements that showed significant associations with AD-related metrics were included in addition to age and education level.

Lastly, in order to determine factors that may have affected group differences in AD-related metrics, a general linear model (GLM) analysis was conducted. In the GLM model, interaction terms (group x MRI R1 & R2* values that were selected from the stepwise regression model) were used in addition to age, group, education level, and individual MRI metal exposure measure.

Statistical significance was defined as α=0.05. Pairwise group comparisons of MRI R1 and R2* measures for each ROI and association analyses were corrected for multiple comparisons using the Stepdown Bonferroni method (Holm 1979) to control the familywise error rate (*FWER*) at *p*=0.05. We reported uncorrected raw *p* values but indicated significant results with *FWER*-correction. We also reported results with p values < 0.09 as a trend level significance. SAS 9.4 was used for all statistical analyses.

## Results

### Demographics and metal exposure characteristics

Welders were older (p=0.048) and had lower education years (p<0.001) than controls. There was no significant difference in UPDRS-III or MoCA scores between welders and controls (p=0.326 and p=0.797; Part I, Table 1). Welders displayed higher short-term (HrsW_90_ and E_90_) and long-term (YrsW and ELT) exposure metrics, and had higher whole blood Mn, Fe, and Pb levels (p’s<0.004; Table 1b).

**Table 1.**
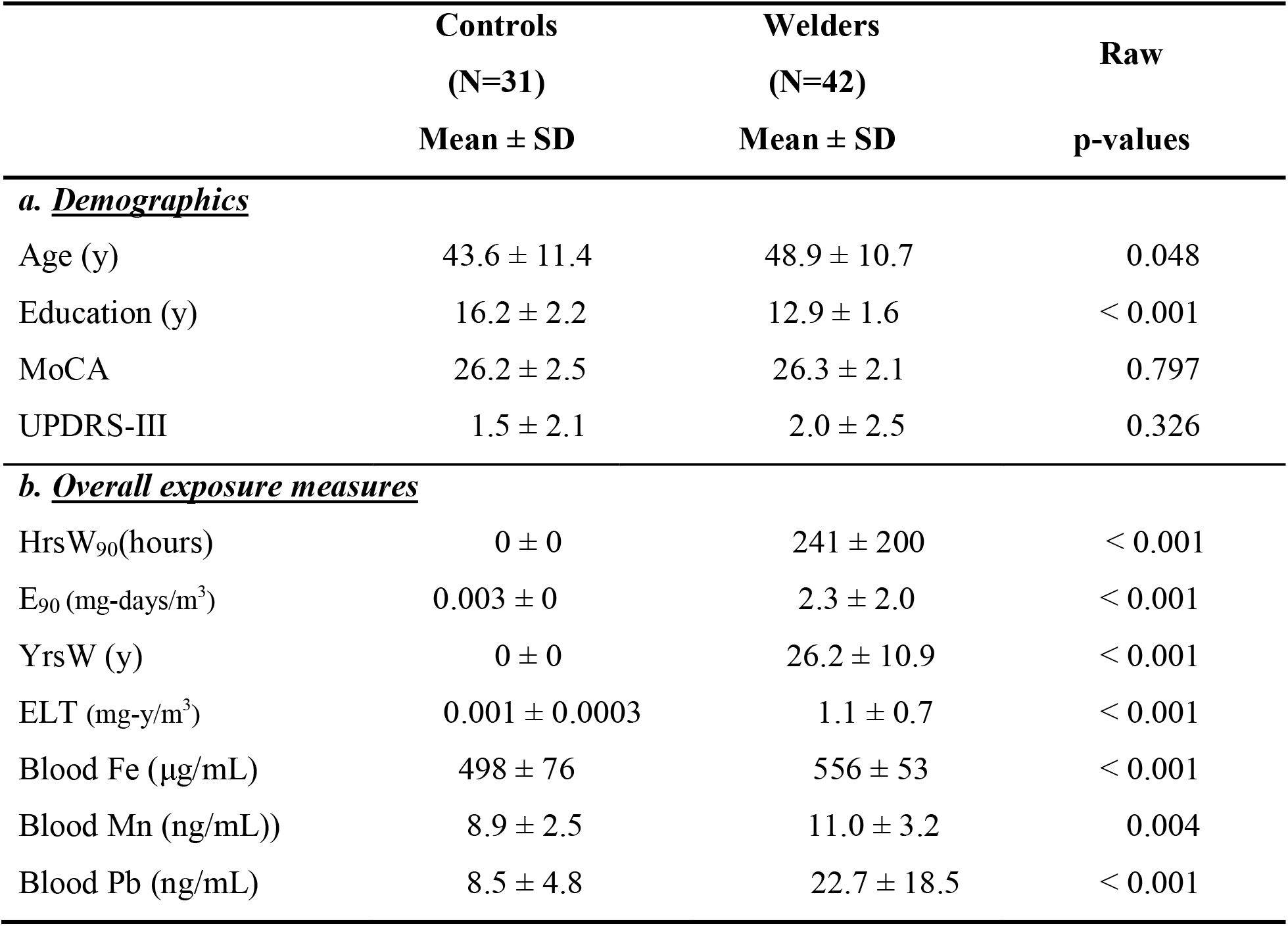
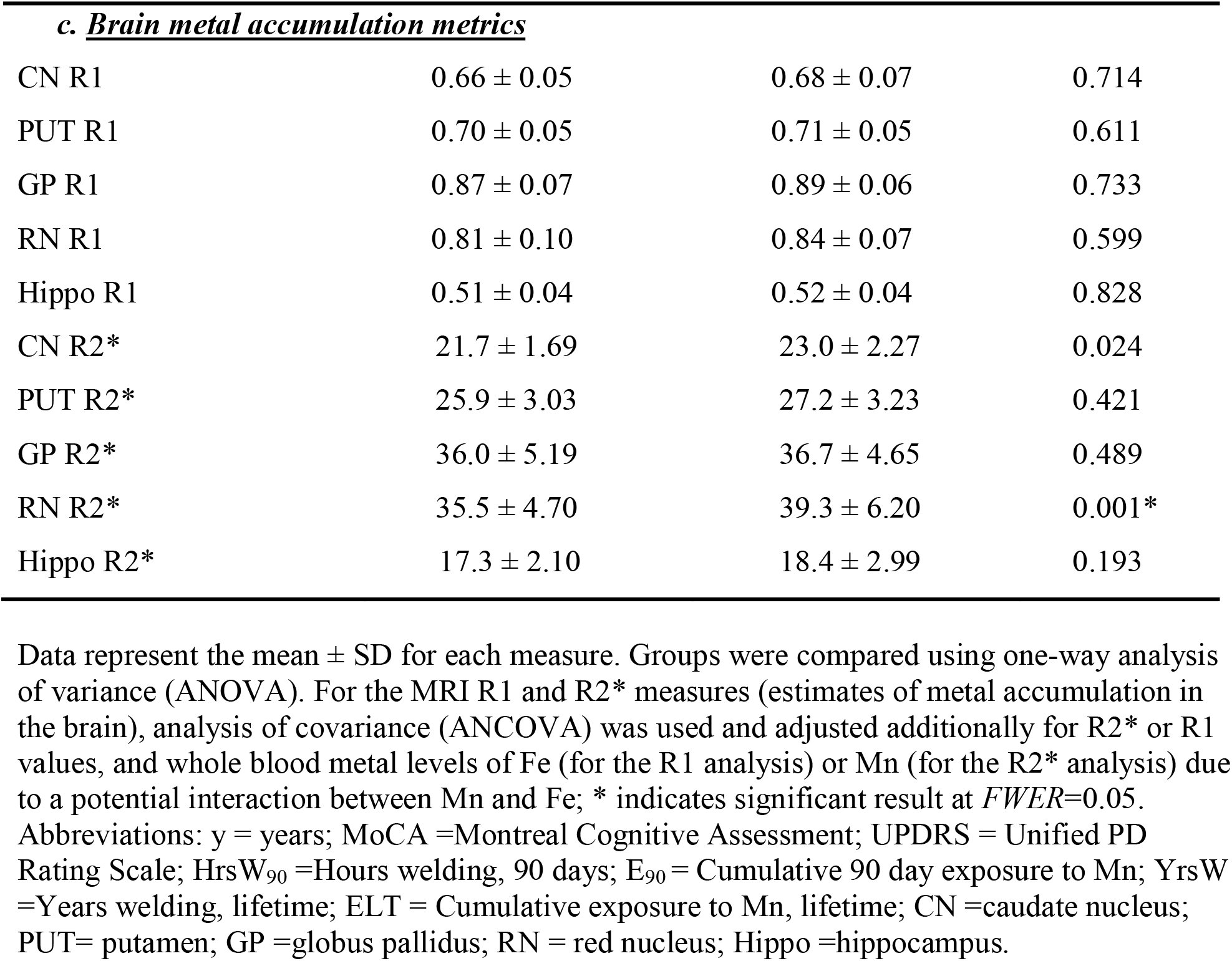
Summary statistics for demographics (a), overall exposure measures (b), and brain metal accumulation metrics (c) for controls and welders.

|  | Controls<br>(N=31) | Welders<br>(N=42) | Raw |
| --- | --- | --- | --- |
| | Mean ± SD | Mean ± SD | p-values |
| ***a. Demographics*** |  |  |  |
| Age (y) | 43.6 ± 11.4 | 48.9 ± 10.7 | 0.048 |
| Education (y) | 16.2 ± 2.2 | 12.9 ± 1.6 | < 0.001 |
| MoCA | 26.2 ± 2.5 | 26.3 ± 2.1 | 0.797 |
| UPDRS-III | 1.5 ± 2.1 | 2.0 ± 2.5 | 0.326 |
| ***b. Overall exposure measures*** |  |  |  |
| HrsW<sub>90</sub>(hours) | 0 ± 0 | 241 ± 200 | < 0.001 |
| E<sub>90</sub> (mg-days/m<sup>3</sup>) | 0.003 ± 0 | 2.3 ± 2.0 | < 0.001 |
| YrsW (y) | 0 ± 0 | 26.2 ± 10.9 | < 0.001 |
| ELT (mg-y/m<sup>3</sup>) | 0.001 ± 0.0003 | 1.1 ± 0.7 | < 0.001 |
| Blood Fe (μg/mL) | 498 ± 76 | 556 ± 53 | < 0.001 |
| Blood Mn (ng/mL)) | 8.9 ± 2.5 | 11.0 ± 3.2 | 0.004 |
| Blood Pb (ng/mL) | 8.5 ± 4.8 | 22.7 ± 18.5 | < 0.001 |
| *c. Brain metal accumulation metrics* |  |  |  |
| CN R1 | 0.66 ± 0.05 | 0.68 ± 0.07 | 0.714 |
| PUT R1 | 0.70 ± 0.05 | 0.71 ± 0.05 | 0.611 |
| GP R1 | 0.87 ± 0.07 | 0.89 ± 0.06 | 0.733 |
| RN R1 | 0.81 ± 0.10 | 0.84 ± 0.07 | 0.599 |
| Hippo R1 | 0.51 ± 0.04 | 0.52 ± 0.04 | 0.828 |
| CN R2* | 21.7 ± 1.69 | 23.0 ± 2.27 | 0.024 |
| PUT R2* | 25.9 ± 3.03 | 27.2 ± 3.23 | 0.421 |
| GP R2* | 36.0 ± 5.19 | 36.7 ± 4.65 | 0.489 |
| RN R2* | 35.5 ± 4.70 | 39.3 ± 6.20 | 0.001* |
| Hippo R2* | 17.3 ± 2.10 | 18.4 ± 2.99 | 0.193 |

Data represent the mean ± SD for each measure. Groups were compared using one-way analysis of variance (ANOVA). For the MRI R1 and R2\* measures (estimates of metal accumulation in the brain), analysis of covariance (ANCOVA) was used and adjusted additionally for R2\* or R1 values, and whole blood metal levels of Fe (for the R1 analysis) or Mn (for the R2\* analysis) due to a potential interaction between Mn and Fe; \* indicates significant result at *FWER*=0.05.
Abbreviations: y = years; MoCA =Montreal Cognitive Assessment; UPDRS = Unified PD Rating Scale; HrsW<sub>90</sub> =Hours welding, 90 days; E<sub>90</sub> = Cumulative 90 day exposure to Mn; YrsW =Years welding, lifetime; ELT = Cumulative exposure to Mn, lifetime; CN =caudate nucleus; PUT= putamen; GP =globus pallidus; RN = red nucleus; Hippo =hippocampus.

<u>For MRI R1measures</u>, there was no significant group difference in any ROI (p’s>0.611). <u>For MRI R2* measures,</u> welders showed significantly higher values in the CN and RN compared to controls (p=0.024 and p=0.001, respectively). The RN R2* comparison remained significant after *FWER* correction. There were no R2* group differences in other ROIs (p’s>0.193; Table 1c).

### Associations of MRI R1 and R2* values with exposure measurements

As previously reported (Lee et al. 2016; Prado-Rico et al. 2022), R1 values in the ROIs were not associated with any blood metal levels within controls (−0.317 < Pearson R’s < 0.309; p’s >0.107) except for a positive correlation between GP R1 and blood Fe at a trend level (Pearson R=0.361, p=0.065). CN R2* values were correlated negatively with blood Mn (Spearman R=-0.428, p=0.029). GP R2* values correlated positively with blood Fe (Pearson R=0.528, p=0.006) and negatively with blood Mn (Spearman R=-0.629, p<0.001) levels. These results remained significant after *FWER* correction. R2* values in the other ROIs did not correlate with any blood metal levels (−0.282 < Pearson R’s < 0.133, p’s>0.163: Supplementary Material 1a).

Within welders, age-adjusted R1 values in the ROIs were not associated with any exposure metrics (Pearson R’s <0.243; p’s >0.121) except for positive associations of PUT R1 and GP R1 with HrsW_90_ and of hippocampal R1 with E_90_ at a trend level Pearson (R’s <0.293; p’s < 0.088). R1 values in the ROIs were not associated with any blood metal levels (−0.180< Pearson R’s <0.127; p’s >0.325). Age-adjusted R2* values in the ROIs did not correlate with any exposure metrics (−0.215 < Pearson R’s < 0.104; p’s >0.189) except for a negative correlation between hippocampal R2* and HrsW_90_ at a trend level (Pearson R=-0.291, p=0.072). CN R2*values were correlated positively with blood Fe (Spearman R=0.360, p=0.043) but not with other blood metals (Pearson R’s <0.108, p’s >0.222). RN R2* values were associated positively with blood Pb (Pearson R=0.507, p=0.003) and negatively with blood Mn (Pearson R=-0.450, p=0.010), but not with blood Fe (Pearson R=-0.057, p=0.758). The RN R2*-blood Pb association remained significant after *FWER* correction. R2* values in other ROIs did not correlate with any blood metal levels (−0.154< Pearson R’s <0.218, p’s>0.231: Supplementary Material 1b).

### Group comparison of AD-related structural and neurobehavioral metrics

As shown in Figure 2, hippocampal volume was similar in both groups after adjustment for age, education level, and TIV (p=0.117; Figure 2a). In contrast, welders had significantly higher hippocampal MD values compared to controls [F (1,69) =6.78, p=0.011; Figure 2b] and lower FA values at a trend level (p=0.066; Figure 2c) after controlling for age and education level.

**Figure 2.**
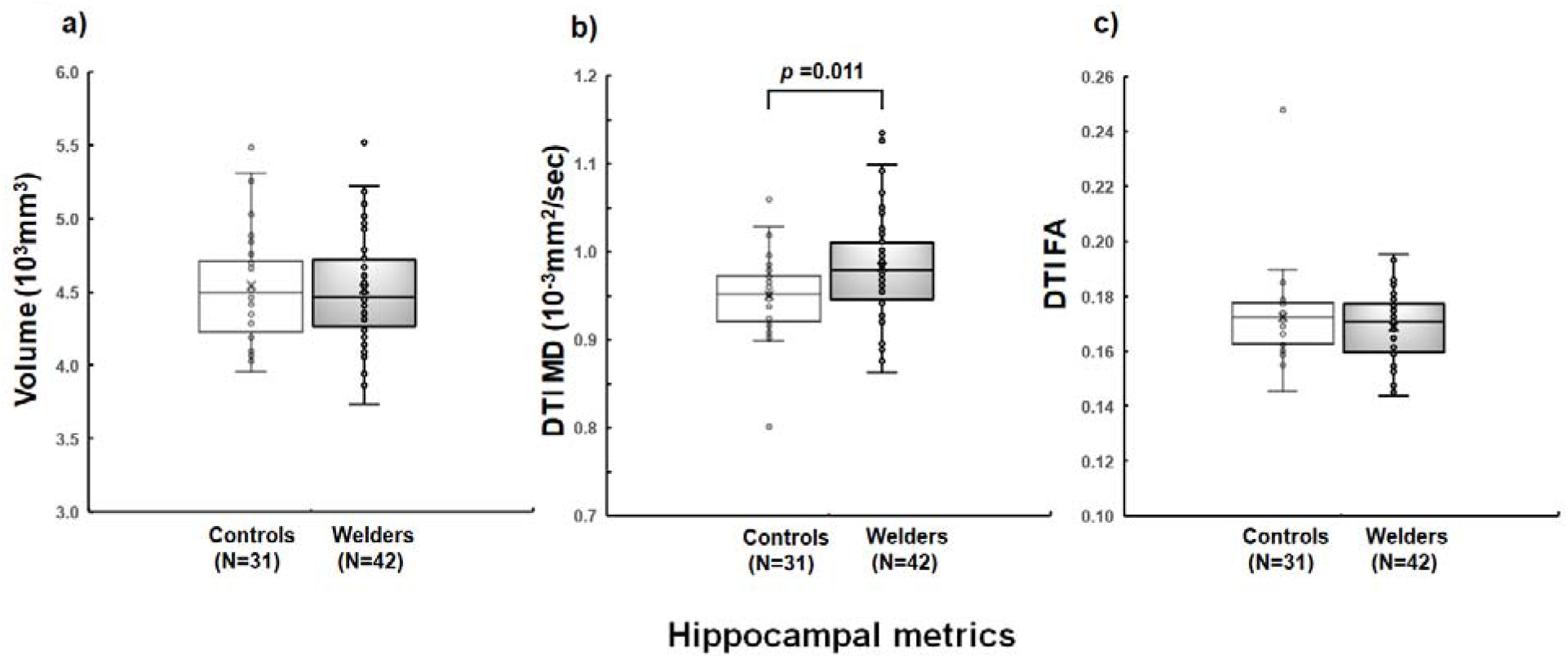
MRI measures in welders and controls. Hippocampal imaging: a) volume b) diffusion tensor imaging (DTI) mean diffusivity (MD), and c) DTI fractional anisotropy (FA).

As shown in Table 2, welders and controls had similar scores in the learning/memory domain (p =0.959). Individual subtest analyses indicated that welders had a lower performance on the delayed *Story Recall* test compared to controls [F (1,70)=4.00, p=0.049] with no group difference in any of the other subtests (p’s >0.192).

**Table 2.** Neuropsychological test results for controls and welders.

|  | **Controls**<br>**(N=31)** | **Welders**<br>**(N=42)** | **Raw** |
| --- | --- | --- | --- |
|  | **Mean ± SD** | **Mean ± SD** | **p-values** |
| ***Learning/Memory*** |  |  |  |
| Mean z-score | -0.26 ± 0.68 | -0.43 ± 0.75 | 0.959 |
| List Learning | -0.64 ± 0.93 | -0.76 ± 1.00 | 0.493 |
| Story Recall<br>(Immediate) | 0.06 ± 0.93 | -0.42 ± 1.04 | 0.151 |
| List Recall<br>(Delayed) | -0.73 ± 0.98 | -0.52 ± 1.01 | 0.192 |
| Story Recall<br>(Delayed) | 0.13 ± 0.90 | -0.46 ± 1.04 | 0.049 |
| Figure Recall<br>(Delayed) | -0.13 ± 1.10 | 0.01 ± 1.16 | 0.261 |

Neuropsychological test results showing the mean ± standard deviation. Standard neuropsychological test scores from the *Repeatable Battery for the Assessment of Neuropsychological Status (RBANS)* were converted to z-scores based on age-adjusted norm scores. Higher z-scores indicate better performance. One-way analysis of covariance (ANCOVA) was conducted with group as the independent variable and the learning/memory domain score and individual subtest scores as the dependent variables with additional adjustment for education level.

### Associations between hippocampal MD and Story Recall metrics

For both controls and welders, there were no significant correlations between hippocampal MD values and *Story Recall* scores (R’s <0.003, p’s >0.932).

### Associations of hippocampal MD and Story Recall metrics with metal exposure

As shown in Table 3a, there were no significant correlations between hippocampal MD values and blood metal levels within controls (Pearson R’s <0.104; p’s > 0.606) except for a negative association with blood Mn at a trend level (Pearson R=-0.370, p=0.057). There also were no significant correlations between hippocampal MD and R1 values in any ROI (Pearson R’s <0.317; p’s >0.107). Hippocampal MD values were correlated positively with CN and GP R2* metrics (Pearson R=0.448, p=0.022 and Spearman R=0.474, p=0.014, respectively) and negatively with hippocampal R2* values at a trend level (Pearson R=-0.366, p=0.066). There were no significant associations of hippocampal MD with R2* values in the other ROIs (Pearson R’s <0.333, p’s>0.256). There were no significant correlations of *Story Recall* test scores with any blood metal levels (−0.145 < Pearson R’s <0.134; p’s > 0.470). There also were no significant correlations of *Story Recall* scores with R1 (Pearson R’s <0.251; p’s >0.206) or R2* (Pearson R’s <0.327, p’s > 0.096) values in any ROI.

**Table 3.** Associations of AD-related hippocampal metrics and Story Recall performance with metal exposure measures for controls and welders.

| **a. Controls (N=31)** |  |  | **b. Welders (N=42)** |  |  |
| --- | --- | --- | --- | --- | --- |
|  | Correlation coefficient R<br>(p-value) |  |  | Correlation coefficient R<br>(p-value) |  |
|  | MD | Story Recall |  | MD | Story Recall |
| HrsW<sub>90</sub> |  |  |  | -0.009<br>(0.957) | -0.113<br>(0.476) |
| E<sub>90</sub> |  |  |  | 0.067<br>(0.673) | 0.025<br>(0.875) |
| YrsW |  |  |  | 0.164<br>(0.300) | 0.181<br>(0.250) |
| ELT |  |  |  | **0.356**<br>**(0.021)** | 0.189<br>(0.230) |
| Fe | 0.104<br>(0.606) | 0.053<br>(0.794) |  | -0.273<br>(0.093) | 0.050<br>(0.761) |
| Mn | -0.370<br>(0.057) | -0.145<br>(0.470) |  | -0.200<br>(0.222) | -0.085<br>(0.607) |
| Pb | -0.014<br>(0.944) | 0.134<br>(0.504) |  | 0.039<br>(0.815) | -0.154<br>(0.348) |
| CN R1 | 0.208<br>(0.297) | 0.241<br>(0.226) |  | -0.013<br>(0.936) | 0.106<br>(0.517) |
| PUT R1 | 0.230<br>(0.248) | 0.188<br>(0.347) |  | 0.105<br>(0.519) | 0.289<br>(0.071) |
| GP R1 | 0.315<br>(0.109) | 0.123<br>(0.531) |  | 0.065<br>(0.690) | 0.127<br>(0.436) |
| RN R1 | -0.082<br>(0.692) | 0.132<br>(0.519) |  | -0.105<br>(0.562) | 0.227<br>(0.204) |
| Hippo R1 | 0.317<br>(0.107) | 0.251<br>(0.206) |  | 0.051<br>(0.756) | 0.131<br>(0.420) |
| CN R2* | **0.448**<br>**(0.022)** | 0.033<br>(0.868) |  | 0.090<br>(0.597) | 0.318<br>(0.055) |
| PUT R2* | 0.333<br>(0.097) | 0.048<br>(0.814) |  | 0.108<br>(0.524) | 0.089<br>(0.560) |
| GP R2* | **0.474**<sup>§</sup><br>**(0.014)** | -0.023<br>(0.910) |  | 0.159<br>(0.346) | -0.040<br>(0.810) |
| RN R2* | 0.232<br>(0.255) | 0.327<br>(0.096) |  | **0.631\***<br>**(<0.001)** | **-0.353**<sup>§</sup><br>**(0.044)** |
| Hippo R2* | -0.366<br>(0.066) | -0.099<br>(0.622) |  | 0.095<br>(0.576) | -0.127<br>(0.453) |

Association analysis results showing Pearson correlation coefficient R (p-value). § indicates Spearman correlation coefficient R (p-value). \* indicates significant result at $FWER=0.05$ . Association analyses were conducted with adjustment for age and education level. For the association analyses with exposure metrics, age-adjusted hippocampal mean diffusivity (MD) and neurobehavioral scores were used. Correlation analyses of hippocampal MD and neurobehavioral scores with blood metal levels of interest were adjusted for the other blood levels in addition to age.

As shown in Figure 3, age-adjusted hippocampal MD values were correlated positively with ELT within welders (Pearson R=0.356, p =0.021; Figure 3a) but not with other exposure metrics (Pearson R’s <0.164, p’s >0.300). There were no significant correlations of hippocampal MD with any blood metal levels (−0.273 < Pearson R’s <0.039; p’s > 0.093). There also were no significant correlations of hippocampal MD with R1 values in any ROI (−0.105< Pearson R’s <0.105; p’s >0.519). Hippocampal MD values were associated significantly with RN R2* values (Pearson R=0.631, p<0.001; Figure 3b). This result remained significant after *FWER* correction. There were no significant associations of hippocampal MD and R2* values in other ROIs (Pearson R’s <0.159, p’s>0.346).

**Figure 3.**
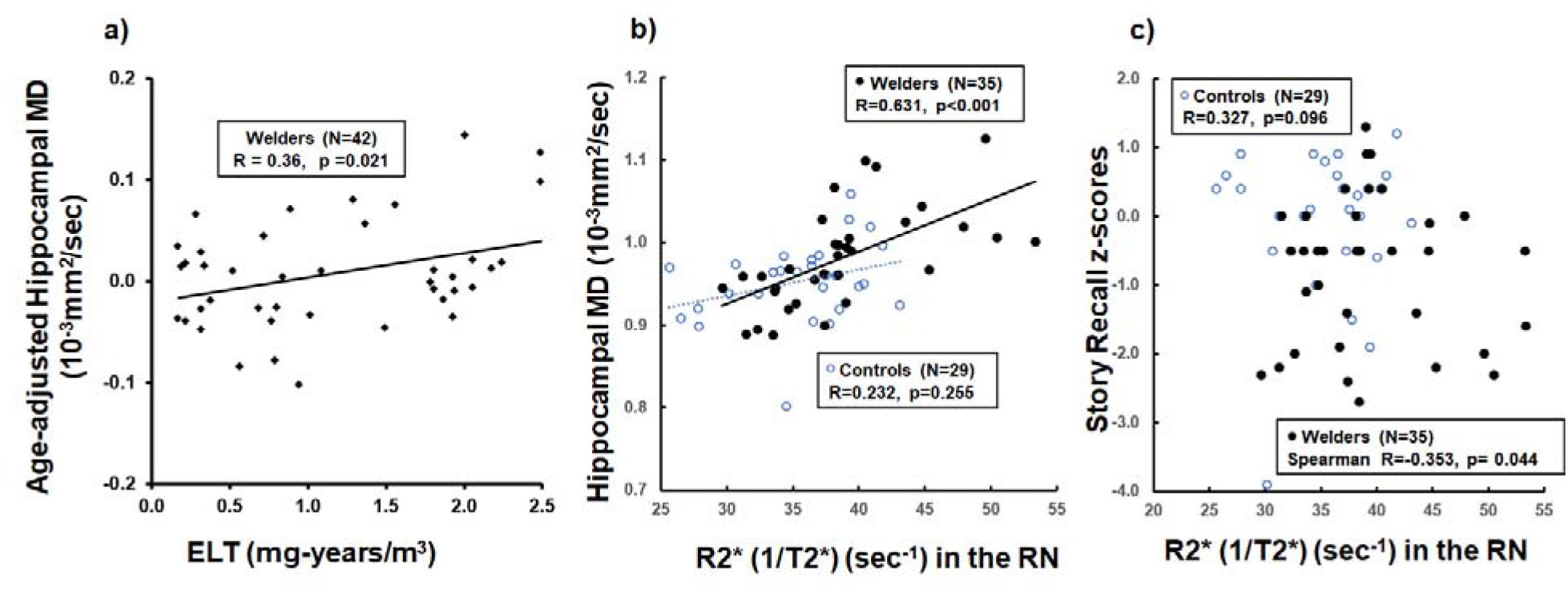
Scatter plots show hippocampal MD values (y-axis) versus a) ELT and b) RN R2* (x-axis); c) *Story Recall* scores (y-axis) versus RN R2* (x-axis) for controls and welders.

There were no associations between *Story Recall* scores and any exposure metrics (Pearson R’s <0.189; p’s > 0.230) or blood metal levels (−0.154 < Pearson R’s <0.050; p’s > 0.348). There also were no significant correlations of *Story Recall* test scores with R1 values (Pearson R’s <0.227; p’s >0.204) except for a positive correlation with PUT R1 at a trend level (Pearson R=0.289, p=0.071). *Story Recall* scores were correlated negatively with RN R2* values (Spearman R=-353, p=0.044; Figure 3c) and positively with CN R2* values at a trend level (Pearson R=0.318, p=0.055), but not in other ROIs (−0.127< Pearson R’s <0.089, p’s > 0.453; Table 3b).

### Stepwise regression analysis to determine factors predicting hippocampal MD and Story Recall performance

For hippocampal MD values, a stepwise regression analysis within controls (using GP R2*, CN R2*, age, and education level as predictors) revealed that GP R2* (partial R^2^=0.334, p=0.002) was a significant predictor. Within welders, a stepwise regression analysis (using ELT, RN R2*, age, and education level as predictors) revealed that RN R2* (partial R^2^=0.376, p=0.001), age (partial R^2^=0.151, p=0.004), and education level (partial R^2^= 0.067, p=0.038) were significant predictors of hippocampal MD. A stepwise regression analysis collapsed across controls and welders (using RN R2*, CN R2*, GP R2*, age, and education level) revealed that RN R2* (partial R^2^=0.301, p<0.001), GP R2* (partial R^2^=0.100, p=0.003), and age (partial R^2^=0.041, p=0.049) were significant predictors of hippocampal MD.

For *Story Recall* performance, stepwise regression analyses were conducted using RN R2*, age, and education level as predictors. Within controls, RN R2* (partial R^2^=0.164, p=0.029) was a significant predictor. Within welders, no variables were significant in the stepwise regression analysis. For the analysis collapsed across controls and welders, education level (partial R^2^= 0.065, p=0.042) was a significant predictor of *Story Recall* performance.

### GLM analysis to determine factors predicting group differences in hippocampal MD and Story Recall performance

For hippocampal MD, a GLM tested the interaction effects of group x RN R2*, group x GP R2*, group x CN R2*, in addition to group, RN R2*, GP R2*, CN R2*, age, and education level. The GLM analysis revealed a significant RN R2* x group interaction effect [F (1,51)=8.02, p=0.007; ß=-0.0020, t=-0.80, p=0.426 for RN R2* in controls; ß=0.0060, t=4.11, p<0.001 for RN R2* in welders]. This result indicates that hippocampal MD values did not change significantly with increasing RN R2* values in controls, whereas they did in welders. This leads to a greater hippocampal MD group difference. The other interaction terms were not significant (F’s <0.57, p’s >0.160).

For *Story Recall* performance, a GLM was conducted using the interaction effects of group x RN R2*, RN R2*, age, and education level. The GLM analysis revealed a significant RN R2* x group interaction effect [F (1,58)=6.16, p=0.016; ß=0.077, t=1.73, p=0.089 for RN R2* in controls; ß=-0.052, t=-1.73, p=0.089 for RN R2* in welders]. This result indicates that *Story Recall* scores tend to increase with increasing RN R2* in controls, whereas they decrease with increasing RN R2* in welders. This leads to a greater group difference on the *Story Recall* test with increasing R2* RN values.

### Post hoc causal mediation analysis

Based on significant findings in the association and GLM analyses, a post-hoc causal mediation analysis (Valeri and VanderWeele 2013; VanderWeele 2015; VanderWeele 2016; Zhang et al. 2016) was conducted to test whether blood Pb affected hippocampal MD or *Story Recall* scores when RN R2* served as a mediator (Figure 4). This analysis was conducted for controls and welders separately, as well as collapsed across both groups with age and education level included as covariates.

**Figure 4.**
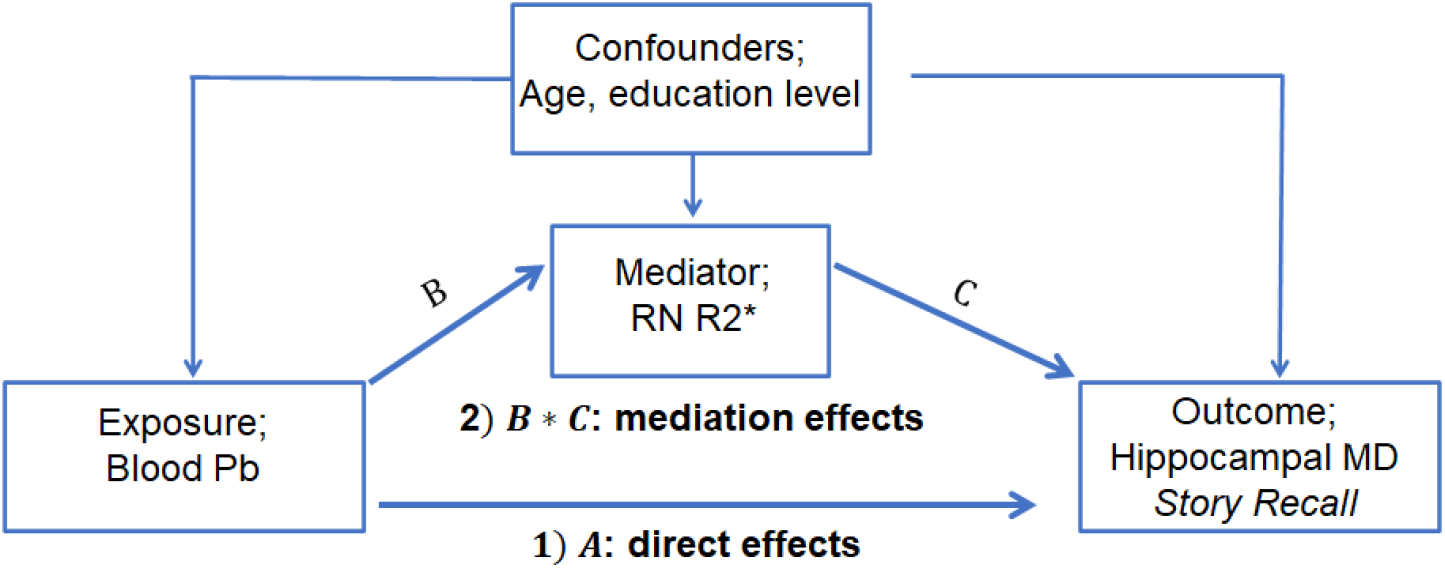
A mediation model to test effects of exposure (blood Pb) on outcome (AD-related metrics: hippocampal MD values or *Story Recall* scores) while RN R2* serves as a mediator after controlling for potential confounders (age and education level): 1) A indicates direct effects of blood Pb on AD-related metrics that are not mediated by RN R2* values; 2) B*C indicates mediation effects of blood Pb on AD-related metrics that are mediated by RN R2* values at the brain level.

For controls, there was no significant direct effect of blood Pb on hippocampal MD (effect that is not mediated by an individual’s RN R2* values; ß=0.00070, z=0.40, p=0.687). The mediation effect of blood Pb on hippocampal MD also was not significant (indirect effect that is mediated by an individual’s RN R2* values; ß=0.00002, z=0.05, p=0.957). Within welders, there was a trend-level mediation effect of blood Pb on hippocampal MD values (ß=0.00038, z=1.70, p=0.089), but the direct effect was not significant (ß=0.00012, z=0.24, p=0.812). Across both groups, there was a significant mediation (ß=0.00043, z=2.10, p=0.036), but not direct (ß=0.00070, z=1.28, p=0.199), effect of blood Pb on hippocampal MD values.

For *Story Recall* scores, there were no significant direct or mediation effects of blood Pb within controls, welders, or across both groups (p’s>0.204).

## Discussion

The current study examined a potential link between metal exposure and AD-related metrics in welders. First, welders exhibited higher R2* values in CN and RN that were associated with higher blood Fe and Pb levels, respectively. Second, welders had higher hippocampal MD and lower *Story Recall* values with no difference in volume or domain-wise learning/memory performance. Most importantly, group differences in hippocampal MD and *Story Recall* metrics were greater with higher RN R2* values. Interestingly, RN R2* values mediated an indirect link between blood Pb and hippocampal MD across both groups. These findings suggest that welding is related to hippocampal microstructural and learning/memory differences, patterns observed in AD-at-risk populations. The AD-related processes may, in part, be linked to Pb exposure that is reflected by higher RN R2* metrics at the brain level.

### Overall metal exposure and brain metal accumulation characteristics in welders

As reported previously, our welders had chronic exposure that was relatively low compared to other studies (Choi et al. 2007; Criswell et al. 2012; Jiang et al. 2008). This low exposure may have contributed to the lack of R1 group differences. The R1 values, however, increased with increasing HrsW_90_ after HrsW_90_ exceeded about half-time welding (data not shown) (Lee et al. 2015b), confirming the R1-metal exposure association.

We also replicated previous findings of higher R2* values in CN and RN in welders (Prado-Rico et al. 2022). R2* values are known to reflect brain Fe accumulation (Peran et al. 2010). Consistently, R2* values in the CN and GP were associated positively with blood Fe levels in welders and controls, respectively. Interestingly, higher RN R2* values in welders were associated with higher blood Pb but not Fe levels, as reported previously (Prado-Rico et al. 2022). Since Pb^2+^ has diamagnetic properties that generate slightly negative magnetic susceptibility (Gaeta et al. 2021), RN R2* values may not reflect brain Pb content per se. Instead, higher RN R2* values may reflect higher brain Fe content secondary to Pb exposure. Consistent with this idea, an animal study found both brain Fe content and R2* values were higher in Pb-exposed rodents (Zhu et al. 2013). Higher R2* values also can reflect pathological processes since substantia nigra R2* is associated with α-synuclein level in parkinsonian autopsy cases (Lewis et al. 2018).

It is important to note that the pattern of brain metal deposition/neurotoxic effects observed in the present study may change if Mn exposure gets higher and starts to accumulate in the brain (Fitsanakis et al. 2011). Consistent with this idea, we observed decreasing RN R2* values with increasing blood Mn levels. Together, the current results suggest that metals other than Mn (e.g., Fe and Pb) may accumulate in the brain when Mn accumulation is not apparent due to low exposure-level. Nevertheless, neuropathologic processes may occur even when there is low-level exposure.

### AD-related hippocampal MD and Story Recall performance differences in welders

As previously reported (Lee et al. 2019), we found higher hippocampal MD without volume differences. DTI markers have been suggested to be more sensitive than hippocampal volume for detecting early AD-related brain changes (Douaud et al. 2013; Kantarci et al. 2005). The exact neuropathological substrates are not clear, but they may reflect microstructural neurodegeneration (e.g., cytoarchitecture and the demyelination process) (Basser and Pierpaoli 2011). This suggests that welding-related microstructural effects may have occurred prior to hippocampal volume differences.

AD-related early neurobehavioral deficits entail learning/memory problems, particularly episodic memory (Bäckman et al. 2001), a system that involves consciously retrieving information that was acquired in a particular time and space (Tulving 1984). In laboratory settings, word-lists or figure recall learning/memory tasks have been utilized to estimate episodic memory ability (Bäckman et al. 2001). In our study, we found no group differences in the learning/memory domain or *List Learning/Recall* and *Figure Recall* subtests. Our welders, however, displayed lower *Story Recall* performance. This result suggests that AD-related early neurobehavioral differences may occur in episodic memory related to story recall before domain-wise performance declines, consistent with previous studies reporting selective episodic memory differences in AD-at-risk-populations (Bäckman et al. 2001; Lim et al. 2014).

The lack of association between hippocampal MD and *Story Recall* is intriguing. Episodic memory is considered a crucial function of the hippocampus (Tulving and Markowitsch 1998). Moreover, hippocampal MD values correlated better with learning/memory performance than did hippocampal volume in elderly subjects (e.g., age >50) (Carlesimo et al. 2010). Our welders’ age, however, ranged from 24-65 years with a mean of ∼50. The standard neuropsychological tests utilized in this study (e.g., *RBANS*) also may not be sensitive enough to detect robustly cognitive differences in asymptomatic populations. Collectively, the current findings of higher hippocampal MD and lower episodic memory metrics prior to differences in volume and domain-wise learning/memory performance are consistent with patterns reported in AD-at-risk populations (Bäckman et al. 2001; Douaud et al. 2013).

### Linking metal exposure to AD-related metrics in welders

Previous welding-related studies focused mainly on neurotoxic effects of excessive Mn exposure in the BG. Recent studies, however, demonstrated Mn accumulation outside the BG (e.g., frontal cortex and hippocampus) (Lee et al. 2015b; Liang et al. 2015). Moreover, metals other than Mn that are abundant in welding fumes (e.g., Fe and Pb) may cause neurotoxic effects. For example, epidemiology studies reported positive associations of Fe or Pb exposure with AD development (Gong et al. 2019; Xu et al. 2018). AD patients had higher hippocampal Fe concentrations that were associated with memory problems (Rodrigue et al. 2013). Pb-exposed workers also revealed hippocampal volume reduction that was associated with blood Pb levels (Jiang et al. 2008).

In our study, higher hippocampal MD values and lower *Story Recall* performance in welders were not associated directly with any blood metal levels, a measure typically representing short-term dynamics of metal exposure. Instead, there was a positive association between hippocampal MD and long-term cumulative welding exposure (ELT metric), suggesting that higher hippocampal MD may result from long-term exposure effects. Most importantly, higher hippocampal MD values and lower *Story Recall* scores were associated with higher RN R2* values. The exact underlying mechanism is not clear. The robust RN R2*-blood Pb association, however, suggests that these AD-related processes in welders may be linked to Pb exposure. Since higher RN R2* values may reflect Pb exposure-related neurotoxic effects, AD-related processes in welders may be due partly to Pb exposure-related neurotoxic effects at the brain level.

A significant mediation effect of blood Pb on hippocampal MD further supports the possibility that the hippocampal microstructural metrics were affected indirectly by Pb exposure and, most interestingly, RN R2* may serve as a mediator at the brain level. It is worthwhile to note that this indirect link between blood Pb and hippocampal MD through RN R2* was significant across both groups but this effect was attenuated to a trend level within welders. The link between blood Pb and *Story Recall* performance also was lacking. It is possible that the sample size of welders was too small to detect a reliable association between blood Pb and AD-related processes. It also is possible that the blood metal level measure may not be sensitive enough to reflect effects of Pb exposure on AD-related differences. Future studies should employ long-term Pb exposure measurements to capture more sensitively the potential links between Pb exposure and AD-related metrics.

The significant associations of R2* values, particularly in the RN, with AD-related metrics also are intriguing. In neural functional networks, the RN is a key region in the cortico-rubral and cerebello-rubral pathways that traditionally have been implicated in motor control (e.g., goal-directed reaching and grasping) (Basile et al. 2021; Cacciola et al. 2019). Recent neuroimaging studies, however, demonstrated that the RN also may be connected structurally and functionally with cortical and subcortical associative areas including the hippocampus (Brockett et al. 2020; Nioche et al. 2009). This implies that the RN may be involved in high-level cognitive functions such as cognitive control (Brockett et al. 2020). The current associations of hippocampal MD and *Story Recall* performance with RN R2* values support the idea that there are RN-hippocampus connections and that these may be involved in high-level cognitive function (e.g., episodic memory).

## Limitations and conclusions

This is the first study suggesting that Pb exposure-related neurotoxic effects in welders may be linked to AD-related processes, with RN R2* values serving as a potential mediator at the brain level. There are several limitations for this study. First, the sample sizes were relatively small. Second, welders had lower education levels that could influence learning/memory task results. Neuropsychological group differences, however, were found only in the *Story Recall* and no other subtests. This argues against the attribution of the *Story Recall* difference to education-related effects. Education level also was used as a covariate. Lastly, blood metal levels were used to assess metal exposure. They, however, are less sensitive in reflecting long-term exposure effects due to metal excretion/elimination over time. Future studies should utilize the best possible long-term metal exposure assessments in order to elucidate causal links between metal exposure and AD-related differences. This may have both scientific and public health relevance.

## Data Availability

All data produced in the present study are available upon reasonable request to the authors

## Acknowledgements

We would like to thank all of the volunteers who participated in this study. In addition, we are indebted to many individuals who helped make this study possible, including: Melissa Santos, Tyler Corson, Lauren Deegan, and Susan Kocher for subject coordination, recruitment, blood sample handling, and data entry; Pam Susi and Pete Stafford of CPWR; Mark Garrett, John Clark, and Joe Jacoby of the International Brotherhood of Boilermakers; Fred Cosenza and all members of the Safety Committee for the Philadelphia Building and Construction Trades Council; Ed McGehean of the Steamfitters Local Union 420; Jim Stewart of the Operating Engineers; Sean Gerie of the Brotherhood of Maintenance of Way Employees Division Teamsters Rail Conference; and Terry Peck of Local 520 Plumbers, Pipefitters and HVAC.

