## supplementary material for "Hippocampal microstructural and neurobehavioral differences in welders are related to higher R2* in the red nucleus"

***Supplementary Material 1. Association analyses of MRI R1 and R2* measures with exposure metrics and blood metal levels for controls and welders***

|  | | a. **Controls (N=31)**   Correlation coefficient R  (p-value) | | | | | | | b. **Welders (N=42)**   Correlation coefficient R  (p-value) | | | | | | | |
| --- | --- | --- | --- | --- | --- | --- | --- | --- | --- | --- | --- | --- | --- | --- | --- | --- |
|  | |  | Fe | | | Mn | | Pb | HrsW_90_ | E_90_ | YrsW | ELT | Fe | | Mn | Pb |
|  | CN  R1 | | | 0.309  (0.116) | -0.255  (0.200) | | 0.024  (0.905) | | 0.056  (0.723) | 0.210  (0.184) | 0.150  (0.344) | 0.141  (0.374) | -0.012  (0.941) | 0.028  (0.867) | | -0.062  (0.709) |
|  | PUT  R1 | | | 0.261  (0.189) | -0.231  (0.246) | | -0.053  (0.795) | | 0.293  (0.060) | 0.243  (0.121) | 0.083  (0.601) | 0.089  (0.576) | -0.065  (0.694) | 0.102  (0.538) | | 0.001  (0.993) |
|  | GP  R1 | | | 0.361  (0.065) | -0.317  (0.107) | | -0.024  (0.907) | | 0.281  (0.072) | 0.083  (0.602) | -0.030  (0.851) | -0.154  (0.331) | -0.161  (0.327) | 0.127  (0.442) | | 0.036  (0.829) |
|  | RN  R1 | | | 0.214  (0.293) | -0.106  (0.608) | | -0.154  (0.452) | | -0.041  (0.816) | -0.174  (0.317) | 0.057  (0.746) | 0.010  (0.959) | -0.067  (0.717) | -0.048  (0.773) | | -0.180  (0.325) |
|  | Hippo R1 | | | 0.193  (0.334) | -0.231  (0.246) | | -0.040  (0.841) | | 0.107  (0.501) | 0.267  (0.088) | 0.068  (0.670) | 0.153  (0.332) | -0.069  (0.677) | -0.048  (0.773) | | -0.046  (0.782) |
|  | CN  R2* | | | 0.053  (0.799) | **-0.428**^§^  **(0.029)** | | 0.053  (0.797) | | -0.132  (0.425) | -0.012  (0.942) | 0.003  (0.988) | 0.003  (0.988) | **0.360**^§^  **(0.043)** | -0.222  (0.222) | | 0.108  (0.556) |
|  | PUT R2* | | | -0.064  (0.757) | -0.282  (0.163) | | 0.004  (0.984) | | -0.107  (0.516) | -0.009  (0.957) | 0.032  (0.847) | 0.032  (0.847) | 0.094  (0.607) | -0.052  (0.776) | | 0.181  (0.322) |
|  | GP  R2* | | | **0.528**^§*^  **(0.006)** | **-0.629**^§^*****  **(<0.001)** | | -0.098  (0.633) | | -0.078  (0.635) | -0.031  (0.853) | -0.075  (0.651) | -0.075  (0.651) | -0.077  (0.674) | 0.093  (0.611) | | -0.124  (0.500) |
| RN R2* | | | | -0.161  (0.433) | -0.225  (0.269) | | -0.204  (0.317) | | -0.064  (0.713) | -0.190  (0.273) | -0.029  (0.869) | 0.104  (0.553) | -0.057  (0.758) | **-0.450***  **(0.010)** | | **0.507***  **(0.003)** |
|  | Hippo R2* | | | -0.061  (0.767) | 0.133  (0.517) | | 0.125  (0.542) | | -0.291  (0.072) | -0.215  (0.189) | -0.173  (0.294) | -0.132  (0.424) | -0.154  (0.399) | 0.051  (0.780) | | 0.218  (0.231) |

Association analysis results showing Pearson correlation coefficient R (p-value). ^§^ indicates Spearman correlation coefficient R (p-value). * indicates significant result at *FWER*=0.05. For the association analyses of MRI R1 and R2* measures with exposure metrics (HrsW_90_, E_90_, YrsW, ELT), age-adjusted R1 and R2* values were used. For the association analyses of MRI R1 and R2* measures with blood metal levels of interest, the analyses were adjusted additionally for the other blood levels.
